# Genomic epidemiology of the early stages of SARS-CoV-2 outbreak in Russia

**DOI:** 10.1101/2020.07.14.20150979

**Authors:** Andrey B. Komissarov, Ksenia R. Safina, Sofya K. Garushyants, Artem V. Fadeev, Mariia V. Sergeeva, Anna A. Ivanova, Daria M. Danilenko, Dmitry Lioznov, Olga V. Shneider, Nikita Shvyrev, Vadim Spirin, Dmitry Glyzin, Vladimir Shchur, Georgii A Bazykin

## Abstract

The ongoing pandemic of SARS-CoV-2 presents novel challenges and opportunities for the use of phylogenetics to understand and control its spread. Here, we analyze the emergence of SARS-CoV-2 in Russia in March and April 2020. Combining phylogeographic analysis with travel history data, we estimate that the sampled viral diversity has originated from 67 closely timed introductions into Russia, mostly in late February to early March. All but one of these introductions came from non-Chinese sources, suggesting that border closure with China has helped delay establishment of SARS-CoV-2 in Russia. These introductions resulted in at least 9 distinct Russian lineages corresponding to domestic transmission. A notable transmission cluster corresponded to a nosocomial outbreak at the Vreden hospital in Saint Petersburg; phylodynamic analysis of this cluster reveals multiple (2-4) introductions each giving rise to a large number of cases, with a high initial effective reproduction number of 3.7 (2.5-5.0).

## Introduction

Since May 11, 2020, Russia is among the four countries with the highest number of confirmed COVID-19 cases ^1^. However, the outbreak in Russia started later than in many neighboring European countries ^2–4^, possibly in part due to early implementation of non-pharmaceutical interventions (NPIs) limiting virus import. Early NPIs included introduction of quarantine for passengers arriving from China on January 23, closing the land border with China on January 31, cancellation of most incoming flights from China on February 1, restricting entrance of non-Russian citizens from China on February 4, and restricting entrance from Iran and South Korea in late February ^5–13^. While the earliest formally confirmed two cases in Russia dated to January and could be associated with direct introduction from China ^14^, no further cases were detected until March 2, 2020, when a woman returning from Italy tested positive ^15^.

Nevertheless, since March 3, a steady increase in confirmed cases has started, with the initial country-wide estimated reproduction number *Rt* of ~2 ^16^. Before March 21, all confirmed Russian cases were imported, while most European countries already had local transmission by this time ^2,17^. Since early March, Russian regional authorities had been implementing their own NPIs. In particular, specific measures were introduced in Moscow and Saint Petersburg, the two largest transportation hubs responsible respectively for 67% ^18–20^ and 10% ^21^ of Russia’s international air traffic. In Moscow, since March 5, all international travellers were temperature checked at the border; and passengers coming from countries with registered cases of SARS-CoV-2 had to report to authorities; while those coming from countries with high case counts at the time, including China, Italy, Spain and the UK, were quarantined ^22^. Since March 14, mandatory quarantine was also applied to passengers’ family members ^23^, and since March 16, it was introduced for all international travellers ^24^. The NPIs at Saint Petersburg were timed similarly ^25^. On March 13, the entrance of non-Russian citizens from Italy was restricted at the state level ^26^, and on March 18, entrance into Russia for all non-Russian citizens for non-emergency reasons was banned ^27^. While inbound flights, mainly returning Russian citizens from abroad, are still operating as of early July, passenger traffic has decreased drastically (e.g., 20-fold at the Moscow Sheremetyevo airport, the one that accepts most international flights during the pandemic ^28,29^).

Here, we report an analysis of 211 SARS-CoV-2 complete genome sequences obtained in Russia between March 11 (when there were just 28 confirmed cases Russia-wide) and April 23 (when there were 62773 confirmed cases) ^30,31^. Phylogenetic analysis reveals distinct introduced lineages associated with transmission within Russia, as well as multiple individual samples phylogenetically intertwined with non-Russian sequences. The largest identified lineage corresponds to an outbreak at the Vreden hospital; phylodynamics analysis of this outbreak reveals between 2 and 4 distinct introductions and initial rapid spread curbed by subsequent establishment of quarantine.

## Results

### Sampling and data acquisition

Samples were obtained from hospitals and out-patient clinics as part of COVID-19 surveillance and sequenced at the Smoroditsev Research Institute of Influenza. We sequenced complete genomes of 135 samples from Russia, including 133 from Saint Petersburg, 1 from the Leningrad region, and 1 from the Republic of Buryatia. Samples were obtained between March 15 and April 23. For analysis, we combined this dataset with additional 76 genomes from Russia available at GISAID ^32^ as of May 26, 2020, obtained between March 11 and April 14. The resulting dataset includes 211 sequences from 25 out of the 85 regions (federal subjects) of Russia (including the Republic of Crimea), with the two regions with the largest numbers of cases, Moscow and Saint Petersburg, most densely covered. Therefore, this dataset is representative of the early outbreak in Russia in terms of geographic spread (Fig. 1a). For phylogenetic context, we also used the 19623 whole-length, high-quality GISAID genomes from the rest of the world available on May 26, 2020.

**Fig 1.**
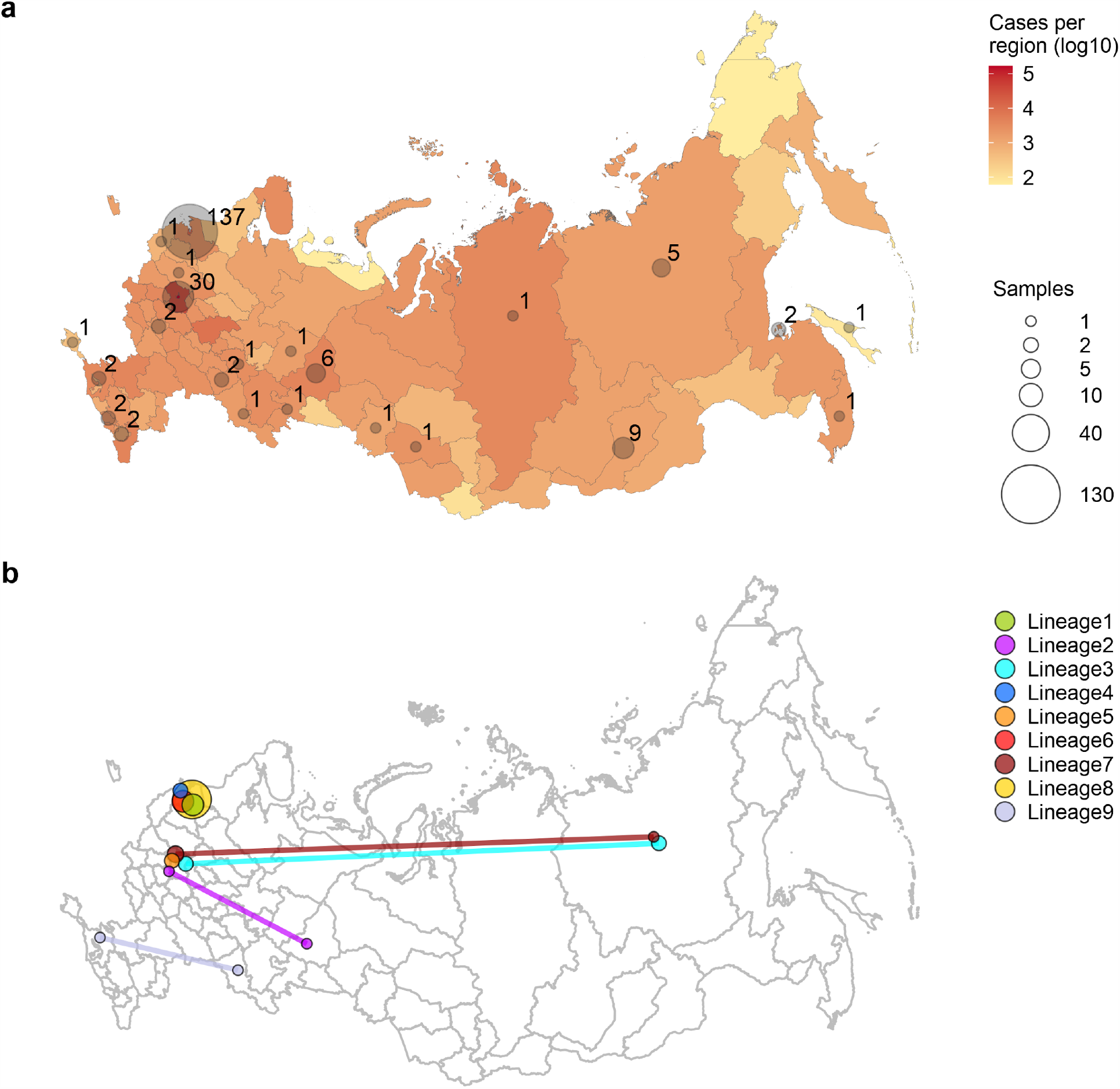
Early epidemiology of SARS-CoV-2 in the Russian Federation. (a) The number of confirmed cases (a, red) and number of sequenced complete genomes (a, grey circles) per region of the Russian Federation (including the Republic of Crimea) as of May 26. Circle sizes are proportional to the numbers of obtained sequences, which are also shown next to the circles. For the purpose of this figure, Moscow was pooled with the surrounding Moscow Region, and Saint Petersburg was pooled with the surrounding Leningrad Region. (b) Identified Russian lineages and corresponding regions; circle size is proportional to the number of sequences belonging to the lineage at this region, and lineages spanning multiple regions are connected by lines.

### Multiple origins of SARS-CoV-2 in Russia

Phylogenetic analysis indicates that the Russian samples are scattered across the SARS-CoV-2 evolutionary tree, representing much of its global diversity (Supplementary Fig. 1). Most samples correspond to the B.1, B.1.1 and B.1.* lineages (PANGOLIN nomenclature ^33^) or clade G, GR and GH (GISAID nomenclature ^34^) which are wide-spread in Europe (Fig. 2). While the predominantly Asian A, B and B.2 lineages comprised 53% of the sampled global viral diversity around the time of Russian border closure (March 27), only 4 (2%) of the Russian samples belonged to them.

**Fig 2.**
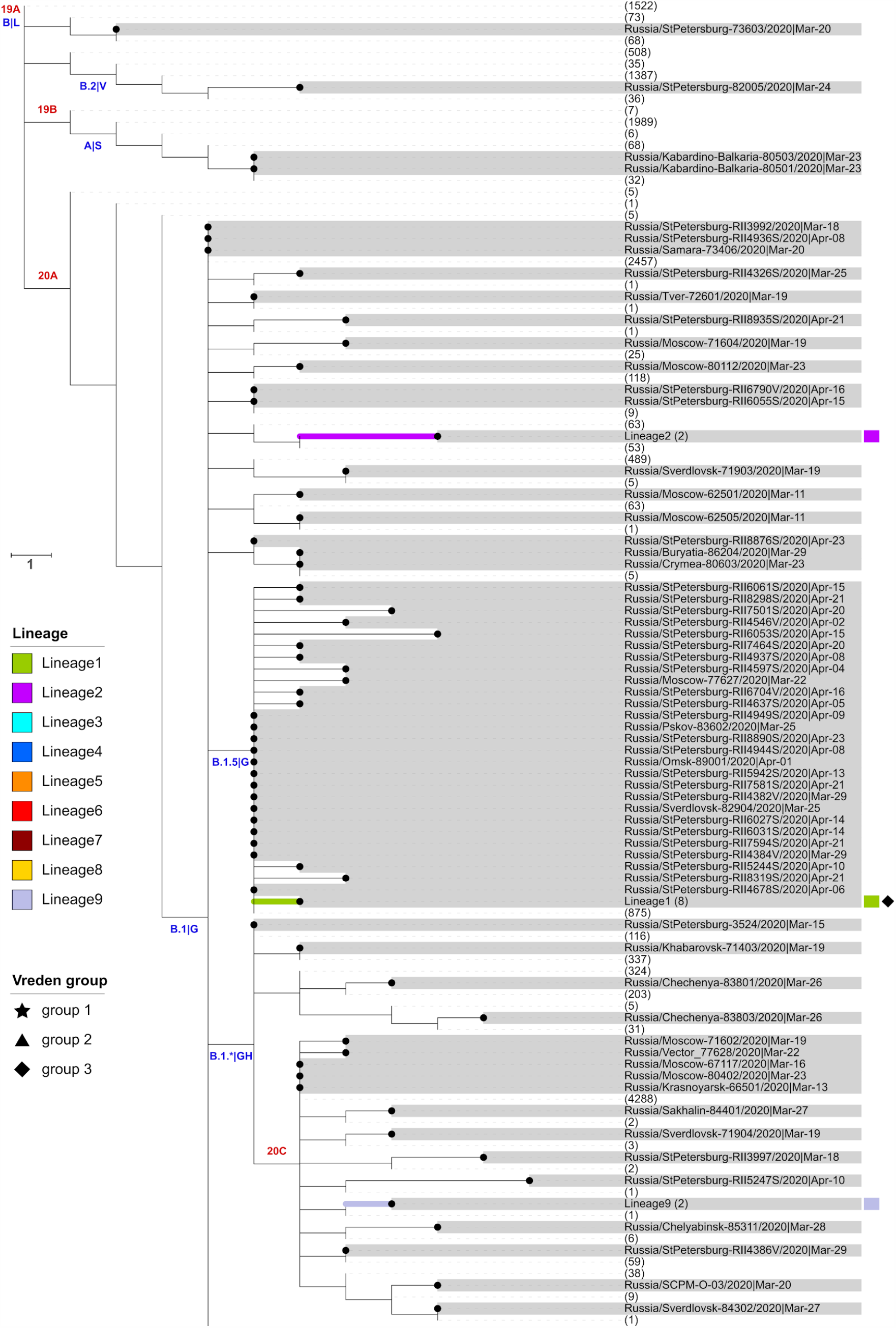
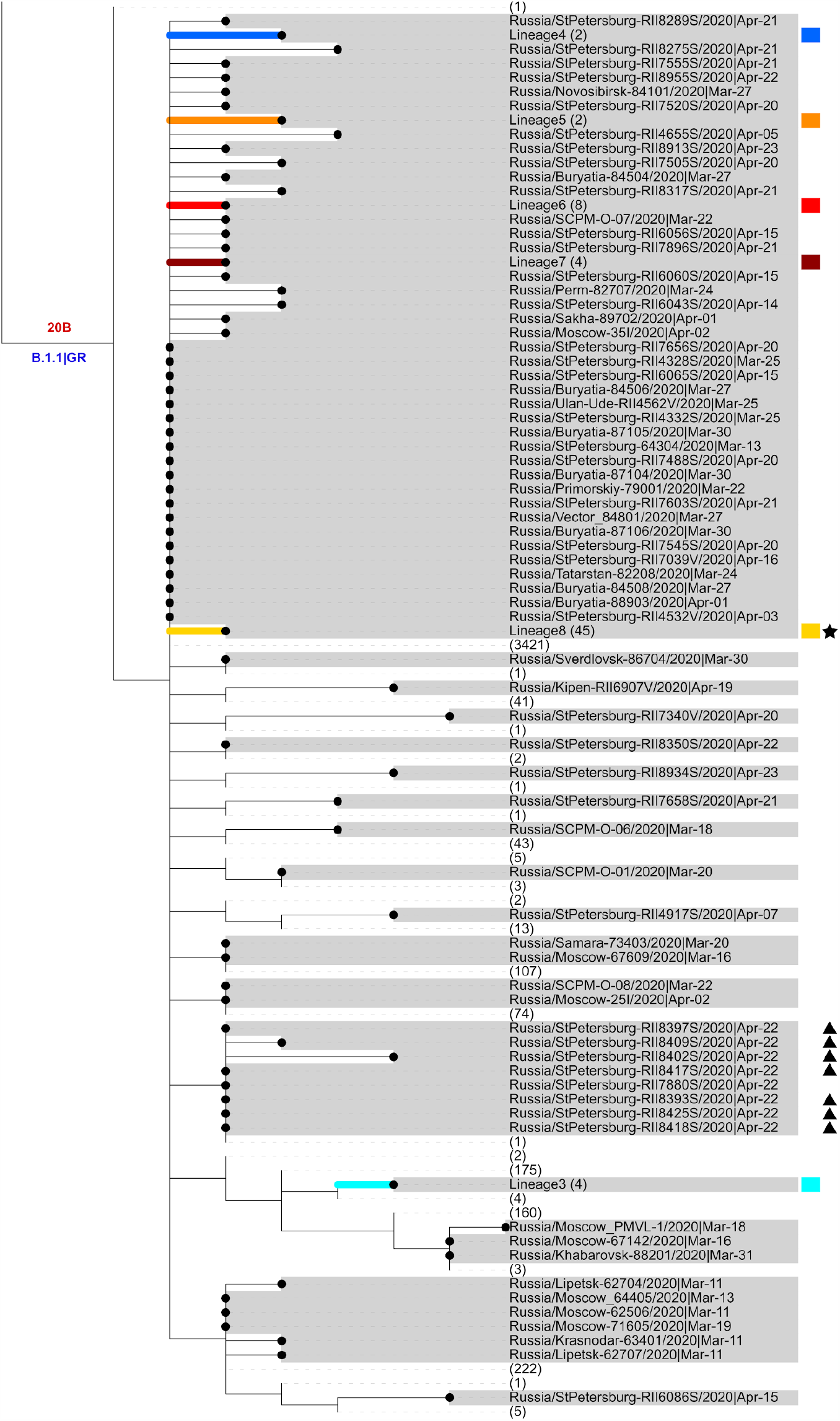
Phylogeny of SARS-CoV-2 in Russia. Russian sequences are identified with dots and highlighted in grey. Russian transmission lineages are truncated to the founder node and highlighted with color (the color scheme is consistent between Figs. 1–3 and 5). Major SARS-CoV-2 lineages are labeled according to Nextstrain ^39^ and PANGOLIN|GISAID nomenclature in red and blue, respectively. Non-Russian sequences and lineages carrying no Russian sequences are truncated, with numbers of such sequences shown in brackets. Sequences from the Vreden hospital and lineages carrying such sequences are marked with star, triangle and diamonds. Branch lengths represent the number of nucleotide substitutions. “hCoV-19/” prefixes are excluded from all sample names for clarity.

We aimed to identify distinct introductions of SARS-CoV-2 into Russia. Phylogenetically, each of the 211 Russian sequences belongs to one of the three categories (Supplementary Fig. 2). Firstly, 77 (36%) of these sequences form the 9 distinct Russian transmission lineages (Fig. 2, 3), defined as monophyletic groups (clades) carrying more than one sequence all of which are Russian. These lineages indicate within-Russia transmission of introduced variants. Five of these lineages had no predating Russian sequences at their ancestral nodes, indicating that they originated from five distinct introduction events (Fig. 3a, c-d; Supplementary Note).

**Fig 3.**
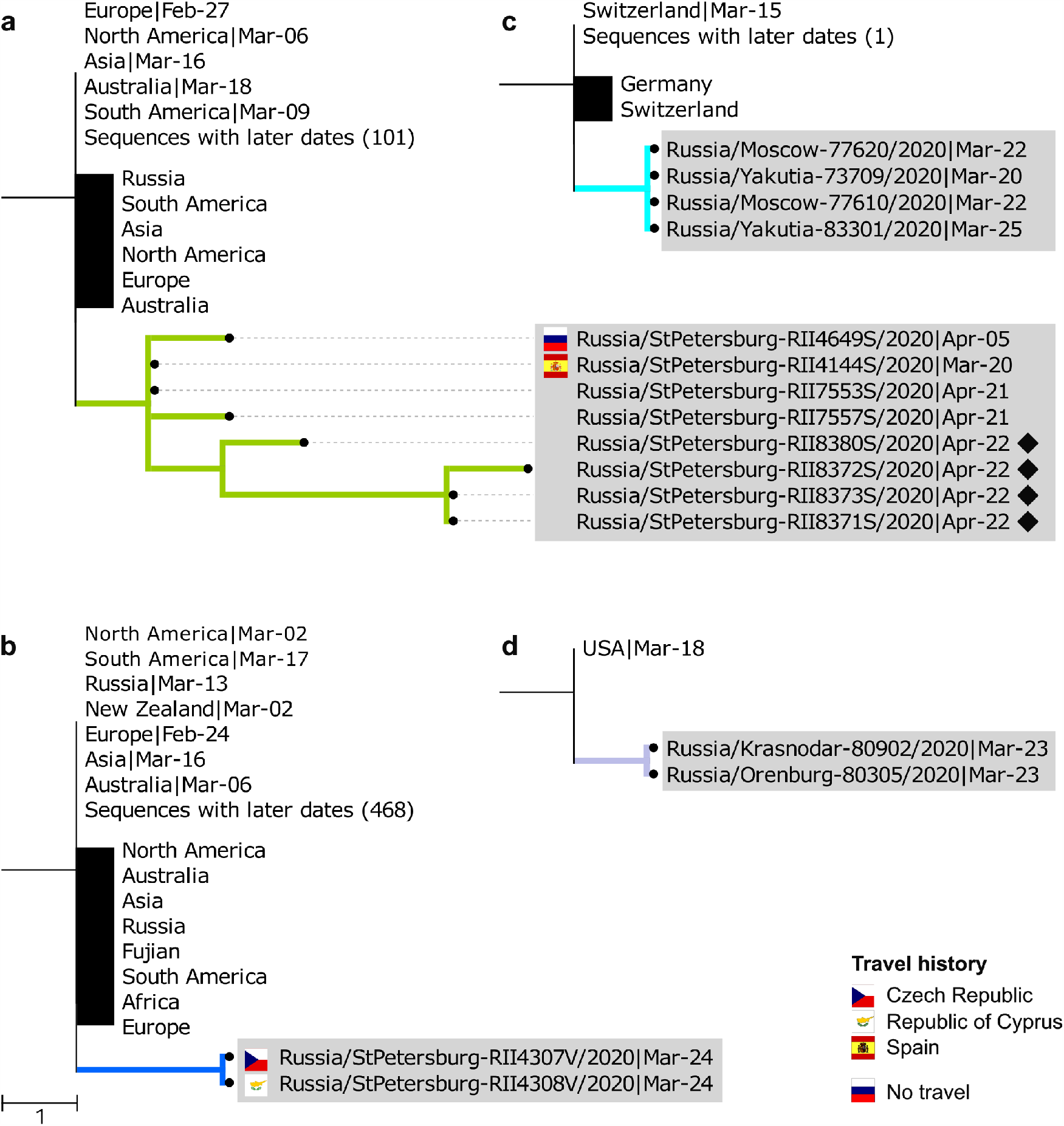
Examples of Russian transmission lineages. Only the phylogeny of the Russian transmission lineage is shown, together with its ancestral phylogenetic node. Russian sequences are marked with dots and highlighted in grey. All other sequences corresponding to an ancestral node (black vertical line) or descendant from it (black rectangle) are truncated, with the region/country and the earliest collection date shown. (a) Lineage 1, a lineage endemic to Saint Petersburg, includes an individual with a history of travel to Spain. (b) Lineage 4 includes individuals with travel history to two different countries, suggesting recurrent introduction. (c) The ancestral node of lineage 3 uniquely maps to Switzerland. (d) The ancestral node of lineage 9 uniquely maps to the USA, and this lineage spans two different regions of Russia. Flags represent individuals with a known history of travel to the corresponding country; the Russian flag shows a known lack of travel history. Diamonds represent samples associated with group 3 of the Vreden hospital outbreak. See Supplementary Fig. 3 for all nine transmission lineages.

The remaining four Russian transmission lineages carried both non-Russian and earlier Russian sequences at their ancestral nodes (Fig. 3b). Such lineages, hereafter referred to as “stem-derived transmission lineages”, could also result from distinct introduction events; alternatively, their last common ancestor could already reside in Russia. To estimate the number of introductions giving rise to the stem-derived lineages, we make use of the direct data on travel history (or lack thereof) available for a fraction of our patients. Using a statistical model, we estimate that these lineages together resulted from roughly one additional introduction event (Supplementary Note). This number could be an underestimate due to undersampling of diversity outside Russia. Indeed, one of the identified lineages (Fig. 3b) involves two samples that had travel history to two different countries, indicating likely double introduction within the same lineage.

Secondly, we observe 73 (34%) singletons each possessing their own characteristic mutations not shared by any other sequences (Fig. 4). These include 33 singletons without any predating Russian ancestral sequences, and 40 singletons stemming from ancestral nodes with earlier Russian sequences (hereafter, “stem-derived singletons”). We assume that the former correspond to sole introduced cases, for a total of 40 such introductions. Most of them had probably not resulted in any within-Russia transmission. However, we find that some of the singleton sequences were sampled from patients without any travel history (Fig. 4c). This indicated that at least some of the singletons likely correspond to distinct introductions that yielded domestic transmission clusters, of which just one representative was sequenced. Using travel data, we estimate that stem-derived singletons resulted from ~6 additional introduction events (Supplementary Note).

**Fig 4.**
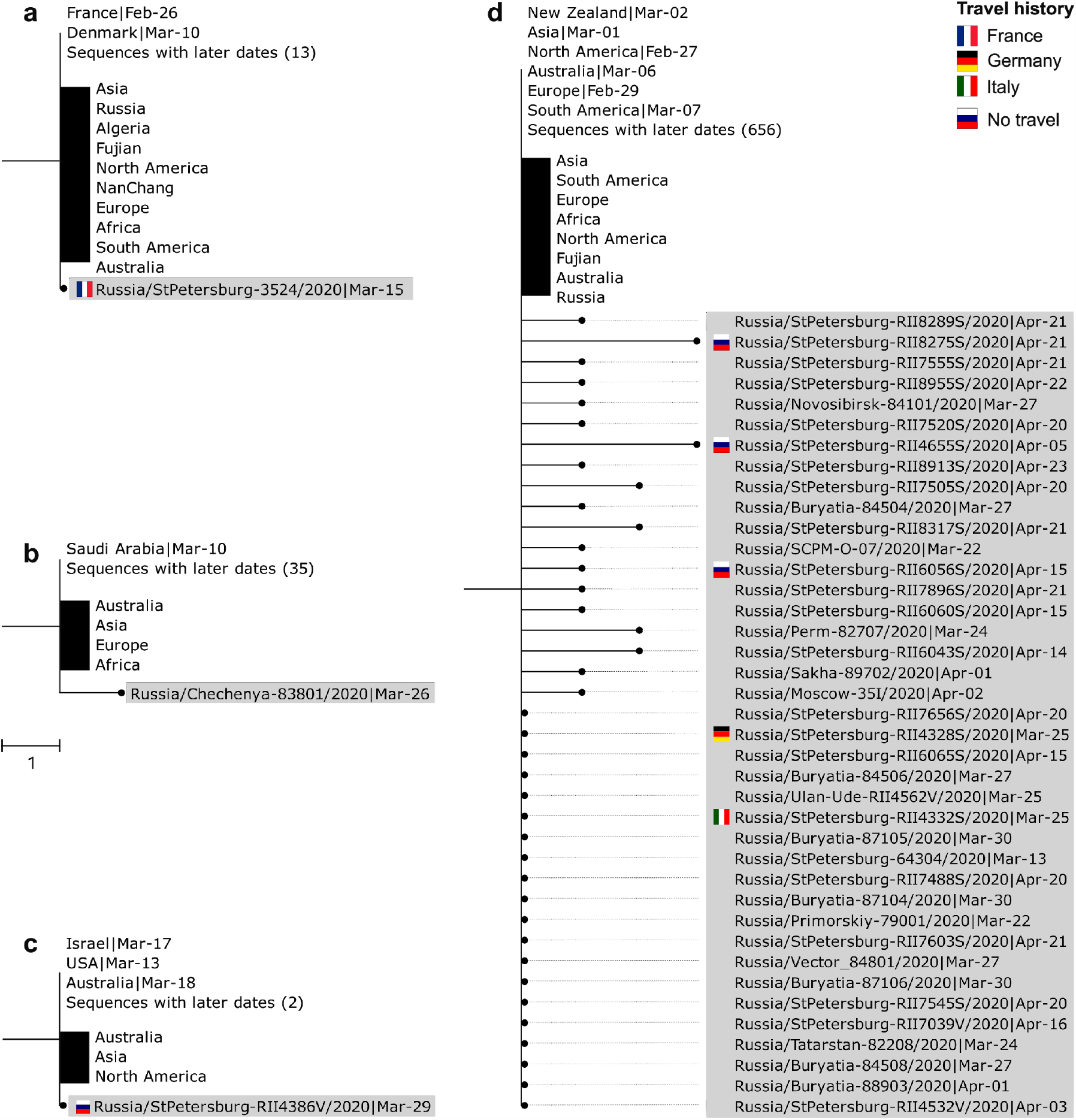
Examples of Russian singletons and stem clusters. Notation is the same as in Fig. 3. (a) The singleton obtained from a patient with known travel history to France has French and Danish sequences at the ancestral node, with French sequences having an earlier date. (b) A sIngleton with a uniquely Saudi Arabian ancestral node. (c) A singleton with known absence of travel history. (d) A stem cluster with associated stem-derived singletons where multiple introductions were observed. See Supplementary Figs. 4 and 5 for all singletons and stem clusters.

Thirdly, the remaining 61 sequences (29%) fell into 12 sets of identical sequences, further referred to as stem clusters, each of which was also identical to some of the non-Russian sequences (Fig. 4d). Again, individual samples within a stem cluster could correspond to distinct introductions or domestic transmission. When data on travel history is available, we find that some of such clusters include multiple individuals with travel history, suggesting that identical sequences were repeatedly introduced into Russia at least in some instances (Fig. 4d). On the other hand, we also observe individuals without travel history, indicating domestic transmission of these variants. From travel data, the estimated number of introductions leading to stem cluster sequences is ~22.

Overall, we estimate the number of independent transmissions into Russia as ~6 resulting in transmission lineages, ~39 resulting in singletons, and ~22 resulting in stem clusters, for a total of 67 events. The uncertainty associated with this estimate is largely dependent on the approach for treating the numbers of introductions leading to stem clusters. If each stem cluster is assumed to originate from exactly one introduction, the estimated number of introductions is 47. If instead each introduction within a stem cluster is distinct, the estimated number of introductions rises to 103.

Phylogenetic analysis indicates that for most Russian transmission lineages, the earliest sample collection dates fall into the range between March 11 and 24, indicating that the corresponding lineages were introduced into Russia not long before (Fig. 5b). Indeed, out of the nine Russian transmission lineages, only two (lineages 6 and 8) had later dates of the oldest sequences. However, those were stem-derived lineages, and the oldest stem sequences corresponding to them dated to March 13, suggesting that these transmission lineages could have also been established by this date. Many (15 out of 33) of the singletons were also collected within this timeframe, although some were collected later (mean date: March 29); together with the fact that many of the singletons have not travelled (Fig. 4, Supplementary Fig. 4), this indicates that they in fact correspond to as yet unsampled transmission lineages. By contrast, most stem-derived singletons were sampled at later dates (mean date: April 7, Mann-Whitney U-test, p=0.014), indicating that they were more likely than non-stem-derived singletons to originate from within-Russian transmission.

**Fig 5.**
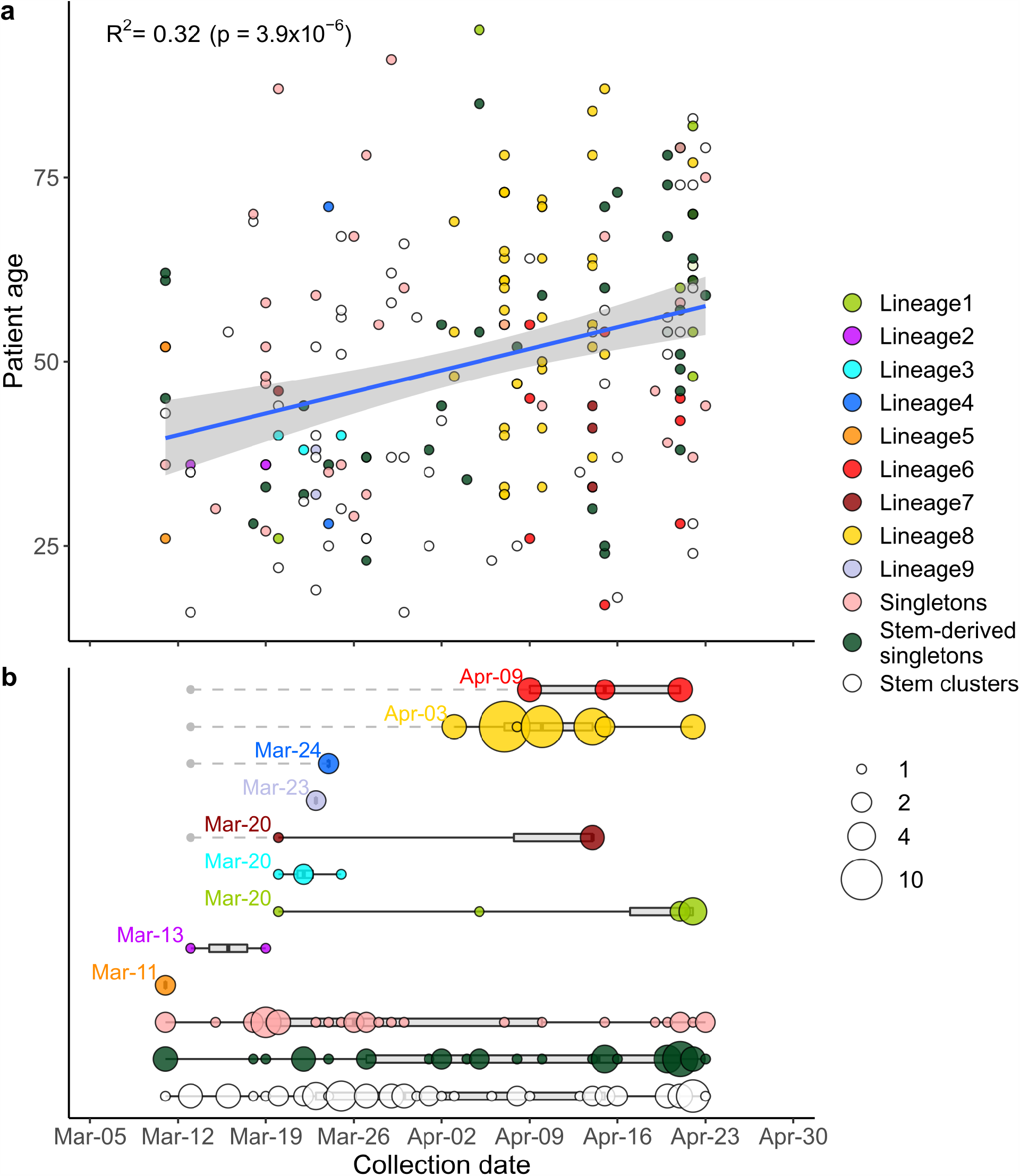
The timeline of SARS-CoV-2 introduction into Russia. Depending on their phylogenetic position, Russian samples are classified as belonging to Russian transmission lineages, singletons or stem clusters (Supplementary Fig. 2). Circles correspond to Russian samples coloured by category. (a) Correlation between sample collection date and patient age. The linear fit is shown (blue line), with the 95% confidence interval indicated as a shaded area. Spearman correlation coefficient is shown. (b) Estimated introduction dates for Russian transmission lineages, singletons and stem clusters. The circle size is proportional to the number of samples. Gray boxes and black lines correspond to the interquartile range and to the full range respectively. For each Russian transmission lineage, the indicated date corresponds to the collection date of the earliest sample. For stem-derived Russian transmission lineages (lineages 4, 6, 7 and 8), the earliest date of the corresponding stem cluster is also shown with a gray dot.

By the time introduction into Russia has started, the virus had already spread through other countries, with the same variant frequently present at multiple locations. Therefore, the source of most introductions could not be established unambiguously. Still, for many of the samples, phylogenetic position is informative of the source. For example, the earliest patient with known travel history has returned to Russia from France, and her sample is nested within a clade with just French and Danish sequences at the ancestral node, with French having earlier dates and therefore a more plausible source (Fig. 4a). For two additional sequences corresponding to regional outbreaks, no direct travel data was available but the probable source could be established from media reports, and was consistent with the phylogenetic position of the corresponding clades. This was the case for the import of clades from Switzerland into Yakutia (the Sakha Republic) (Fig. 3c) ^35,36^ and from Saudi Arabia to the Chechen Republic (Fig. 4b) ^37^.

Overall, out of the 13 patients with known travel history (11 direct + 2 from media reports), the country of origin is consistent with the sampling locations of the same or ancestral nodes in 9 cases, including the 3 cases when it is uniquely identified. In one case (Supplementary Fig. 5b), the travel direction (Egypt) is inconsistent with the phylogenetic position of the sample, and in the remaining three cases, there is not enough phylogeographic data to make a call. For the same 9 out of the 13 patients, we were able to correctly and uniquely identify the source continent (Europe in all cases).

Consistency between direct travel history and phylogenetic position motivated us to attempt to infer the sources of Russian samples phylogeographically. In the absence of travel data, we can position the source of one transmission lineage (lineage 9, Fig. 3d) as the USA; and of five singletons, to Denmark (Supplementary Fig. 4c, l), France (Supplementary Fig. 4u), Chile (Supplementary Fig. 4y), and England (Supplementary Fig. 4ab). For 6 additional singletons, we are able to position the source to the continent (Europe in all cases). Finally, we infer the origin of one stem cluster as Sweden (Supplementary Fig. 5c), and of two more stem clusters, as Europe.

The individuals importing the virus and seeding the Russian transmission lineages were not a random sample of the population. Very early samples were collected from patients who were on average younger than those sampled later (Fig. 5a). This is consistent with the major role of younger Russians in the import of virus into Russia ^38^, possibly because they comprised a larger share among the people returning from business trips or holidays.

No non-Russian sequences were nested within predominantly Russian clades (Figs. 2, 3, Supplementary Fig. 1). Therefore, we observe no sign of export of SARS-CoV-2 outside of Russia.

### Temporal dynamics of SARS-CoV-2 spread in Russia

Following introduction, the virus has spread throughout Russia. Four out of the 9 identified Russian transmission lineages, and 8 out of the 12 stem clusters, span multiple regions (Figs. 1b, 3c-d, 4d). As Moscow and Saint Petersburg are major transport hubs, together responsible for 77% of the international air traffic in Russia, we hypothesized that the virus was introduced through these cities, and spread throughout Russia from them. Contrary to this hypothesis, among the Russian stem clusters and singletons, the samples from Moscow or Saint Petersburg do not sit on shorter branches than samples from other regions; in fact, branches leading to them tend to be slightly longer (0.88 vs. 0.37, p=0.006, permutation test), probably because of more extensive regional sampling early in the outbreak. Thus, we see no evidence for a preferential direction of transmission within Russia, suggesting that the Russian epidemic has been seeded by near-concurrent introduction into multiple regions.

### Vreden hospital outbreak

A major transmission cluster corresponded to the nosocomial outbreak at the Vreden Russian Research Institute of Traumatology and Orthopedics in Saint Petersburg (hereafter, the Vreden hospital) ^40,41^. According to an internal investigation, the suspected zero patient at the hospital had surgery on March 27. While routine COVID-19 testing at the Vreden hospital began on March 18, the earliest samples that tested positive were obtained on April 3. Quarantine was gradually introduced between April 7 and April 9, which involved a complete lockdown of the hospital, isolation of units from each other, and shutdown of the hospital-wide ventilation system. 474 patients and 270 medical workers remained inside the hospital for the following 35 days.

Our dataset contains SARS-CoV-2 genomes obtained from 52 of the Vreden hospital patients or medical workers. Phylogenetic analysis indicates that these samples form three distinct groups, each defined by its own set of mutations. The largest group, group 1, includes 41 sequences obtained between April 3 and April 22 and represents a distinct Russian transmission lineage (lineage 8, star in Fig. 2). This lineage derives from a very prolific ancestral node which has seeded multiple lineages throughout the world, including five of the Russian transmission lineages, so its origin cannot be positioned phylogeographically. Group 2 contains 7 out of 9 sequences in another clade, which also carries one non-Russian (English) sequence (triangles in Fig. 2). Finally, group 3 includes 4 sequences and represents a clade of its own within another Russian transmission lineage (lineage 1, diamonds in Figs. 2 and 3a). While samples from group 1 came from different units located at different floors of the Vreden hospital, samples from groups 2 and 3 each came from its own unit.

Groups 1 and 2 are phylogenetically remote from group 3, with six mutations separating the most recent common ancestors (MRCAs) of groups 1 and 2 from group 3 (Fig. 2). Groups 1 and 2 belong to the B.1.1 lineage defined by three mutations at positions 28881, 28882 and 28883, and are further defined by mutations at positions 26750 and 1191, respectively. By contrast, group 3 belongs to the B.1.5 lineage, and is supported by the mutation at position 20268 which is widespread over the world and appeared early in phylogenetic history, as well as by two additional mutations. This provides strong evidence that group 3 originates from a separate introduction to that of groups 1 and 2.

To understand the spread of the outbreak at the Vreden hospital, we performed a Bayesian phylodynamic analysis using the birth-death skyline model ^42^ of BEAST2 ^43^. Given the possibility of multiple introductions, we analyzed the whole Vreden dataset comprising groups 1, 2 and 3; and also its two subsets consisting of groups 1 and 2, and just of group 1. The results are summarized in Figs. 6–7 and Supplementary Tables 4, 5 and 6.

**Fig 6.**
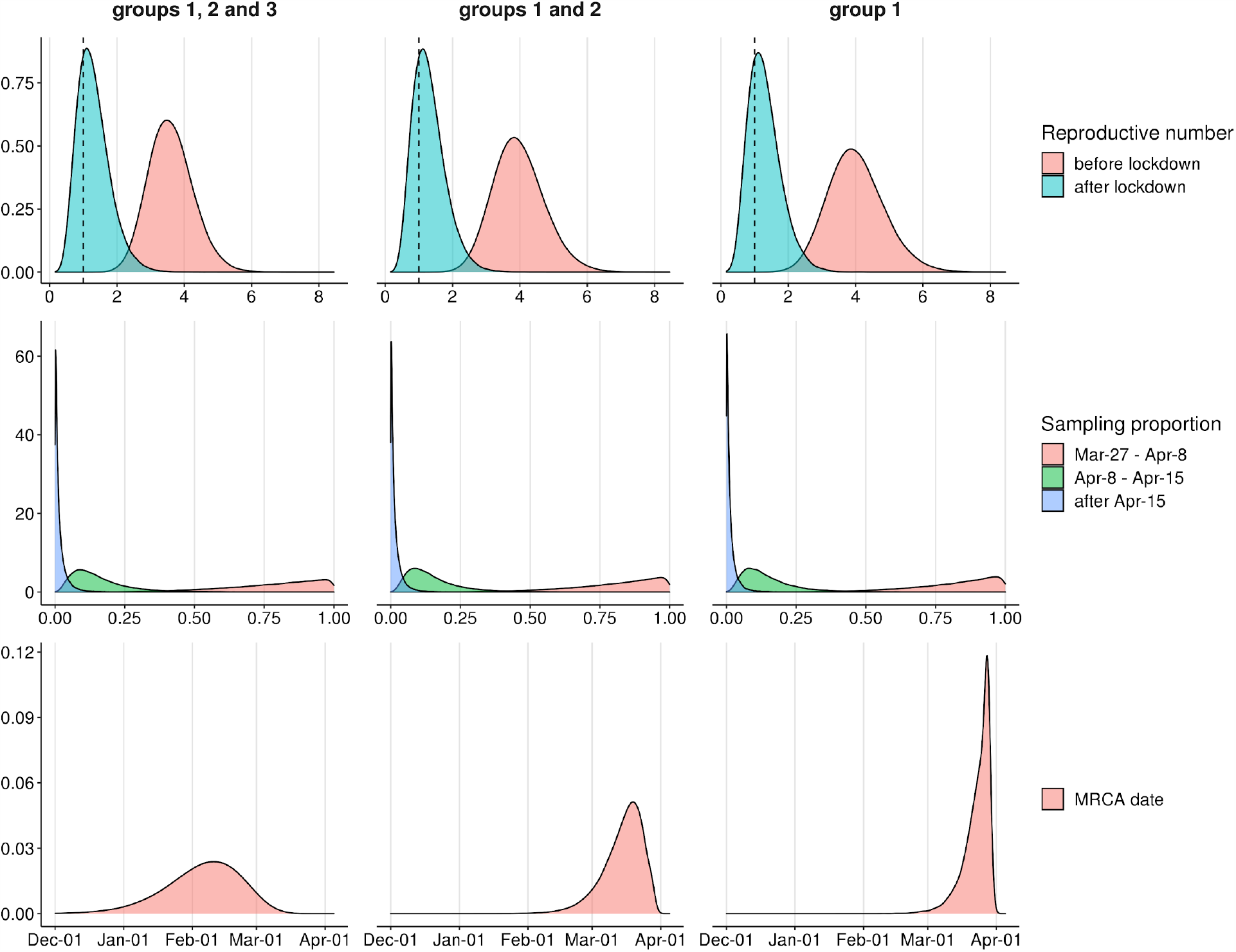
Vreden hospital outbreak parameter estimates produced by birth-death skyline model in BEAST2. Panels show posterior distributions of effective reproductive number Re (upper panel) with the dashed vertical line corresponding to Re=1, sampling proportion (middle panel) and the date of the MRCA (lower panel) for analyses based on Vreden hospital samples from groups 1, 2 and 3 (left column), groups 1 and 2 (middle column) and group 1 (right column).

We found that the Bayesian analysis supports at least two distinct introductions of SARS-CoV-2 into the Vreden hospital. This is based on the deep split between group 3 and groups 1-2. The MRCA of all three groups dates to February 4 (95% CI January 1 - March 7). This is almost two months prior to the assumed date of introduction (March 27), implying that group 3 and the remaining Vreden samples were introduced independently.

A third introduction into the Vreden hospital is also highly probable. Indeed, the MRCA of groups 1 and 2 dates to March 15 (95% CI February 25 - March 31). As there was no sign of infection at the hospital before the end of March, it is quite likely that these two groups originated through separate introductions. The root of the group 1 dates to March 23 (95% CI March 11 - March 30), which is consistent with the suspected illness period of the zero patient. Additional evidence that groups 1 and 2 originate from distinct introductions is provided by the fact that the clade that includes group 2 also carries a non-Russian (English) sequence (Fig. 2).

Finally, a fourth introduction is also possible, suggested by the deep split within the group 3 (Fig. 7). The MRCA of this group 3 dates to March 17, although this estimate has a broad confidence interval which overlaps the supposed illness period of the zero patient.

**Fig 7.**
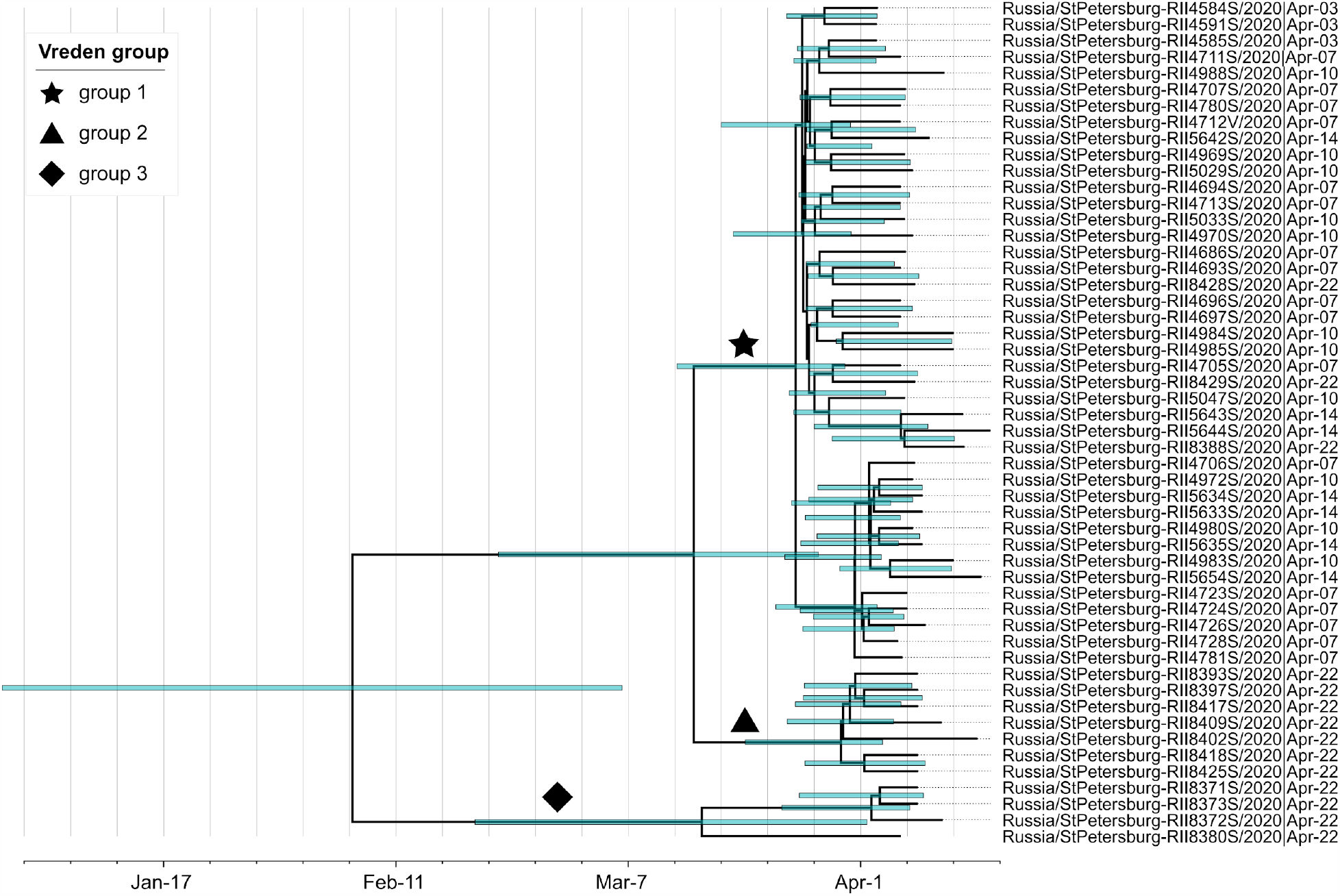
Maximum clade credibility tree for the Vreden hospital outbreak. Groups 1, 2 and 3 are marked by a star, a triangle and a diamond, respectively. Green bars represent 95% credible intervals.

We estimated the phylodynamic parameters before and after the quarantine measures were introduced. In all three analyzes, the estimates of phylodynamic parameters were stable and consistent with each other. We found that the effective reproductive number Re was 3.72 (95% CI 2.48-5.05) before April 8, and dropped to 1.38 (95% CI 0.48-2.41) after April 8 (Fig. 6). Remarkably, the credible intervals for these estimates do not overlap, indicating a statistically significant slow down of the outbreak. The sampling proportion decreased with time, indicating that the number of sequenced samples did not keep up with the rapidly growing case counts (Fig. 6, Supplementary Table 3).

## Discussion

The ongoing pandemic of SARS-CoV-2 has involved rapid spread of the virus across the borders of most nations within the few weeks of February and March. While Russia was behind many of the neighboring countries in the initial rise of the case counts, it has rapidly caught up in the following weeks. By analyzing the phylogenetic distribution of 211 early COVID samples from those dates, we provide details of this process, shedding light of the patterns of transborder transmission of the virus and the factors that affect it.

SARS-CoV-2 accumulates substitutions at the average rate of ~1 per 1000 nucleotides per site per year ^44^, which means that its genome accumulates on average just one mutation per 2-3 transmissions. Therefore, phylogenetic trees have lower resolution than transmission trees, meaning that transmission history cannot be fully resolved from phylogenetics alone. In particular, in the absence of complete data on travel history, there is no simple rule for counting the number of introductions. A common rule of thumb is counting the number of country-specific clades ^45–49^. However, multiple introductions can result in a single clade if the viral diversity abroad is undersampled ^50–52^; and a single introduction can result in multiple clades if their last common ancestor has already been introduced ^47^.

By using direct travel data, we show that both these problems hold. Indeed, we find transmission lineages apparently co-introduced from multiple countries (Fig. 3b) or singletons without any history of travel (Fig. 4c). The uncertainty in the number of introduction events is the highest for identical sequences with broad geographic distribution, e.g., the last common ancestor of lineage B.1.1. This node constitutes a stem cluster of 100 identical Russian sequences, as well as 4323 sequences from outside Russia. It is the immediate ancestor to five Russian transmission lineages and 19 stem-derived Russian singletons, so how many times this sequence has been introduced into Russia strongly affects the overall counts of the number of introductions. Travel data indicate that a stem group can carry a combination of multiple introduced and domestically transmitted sequences, complicating the inference of the number of introductions.

Under a simple statistical model combining genetic and available travel data, we estimate that the sampled diversity of SARS-CoV-2 in Russia originated from 67 introductions. Since this corresponds to roughly one introduction per each three sequences sampled, the actual number of introductions was probably much higher, and its estimate will likely increase as more sequences are sampled.

Contrary to some previous reports ^47,53^, we find that the phylogeographic position of a lineage is predictive of its origin. For some of the Russian samples, we are able to infer unambiguously or with little ambiguity their origin, and when direct travel data is available, it supports our claims. The difference from the UK study may be due to the fact that most Russian lineges originated late, when the source European lineages were already well established. Still, for many of the introductions, phylogeography is not informative of their origin; moreover, phylogeographic inferences are expected to be biased in the presence of uneven sampling between countries. This illustrates the need of combining multiple data sources, including direct travel data, for understanding viral origin and spread ^54^.

Although it is hard to ascribe epidemiological results to specific NPIs, our analysis suggests that the border closure with China implemented in February has effectively curbed the virus introduction into Russia from the Asian direction. Indeed, only four of our samples belong to lineages A, B and B.2 (GISAID clades S, L and V, respectively), which predominantly originated in Asia; and two of those sequences are nested within other European subclades, indicating that the import was through Europe. This fraction is not representative of global case counts at that time, and is instead reflective of travel patterns and history of border closures. It is also in contrast to the situation in other countries where the outbreaks started earlier, and were probably seeded by direct introduction from Asia ^55–57^.

As a result, the major source of the SARS-CoV-2 introductions into Russia was Europe. We see robust evidence for transmission within Russia, but no evidence of export of SARS-CoV-2 outside Russia. This is consistent with a lower number of international travelers leaving Russia compared to the number returning to Russia after the start of the pandemic; later start of the outbreak compared to that in neighboring countries; and/or higher efficiency of border closure at later stages of the outbreak. More generally, this is consistent with Europe being a global source, rather than a sink, of infections up to early March ^58^.

For most of the discovered transmission lineages, the earliest sampled sequence was collected between March 11 and 24 (Fig. 5b). In the larger UK dataset, the mean time between the importation date of a lineage and its earliest sampling date within the UK was estimated to be approximately two weeks, although this depends on many factors including lineage size and sampling intensity ^47^. If this can be extrapolated to Russian transmission lineages, this implies that these lineages typically originated from imports in the last week of February and the first week of March. A contributing factor could have been intensive travel around the Russian state-mandated long holidays of February 22-24 and March 7-9. Further establishment of Russian transmission lineages could be limited by NPIs, in particular, by introduction of mandatory quarantine for incoming travelers on March 5, as well as by the overall radical reduction in international travel after these dates.

Detailed analysis of localized transmission clusters helps understand viral spread. Well-studied examples include the Diamond Princess cruise ship ^59–62^; the Grand Princess cruise ship ^63^; an international conference in Boston ^46^; a community living facility in the Boston area ^46^; and the nosocomial outbreak in the Netcare St. Augustine’s Hospital in South Africa^64^. In all but one of these cases, the outbreaks were genetically homogenous, indicating that they each arose from a single case. In the community living facility, multiple introductions have occurred, but there was a dominant clade that included nearly all the samples, while other clades were rare ^46^. By contrast, at the Vreden hospital outbreak, we observe multiple (2-4) introductions, each of which gave a prolific clade. This indicates that this outbreak could have originated from multiple superspreading events. Furthermore, we estimate the initial effective reproductive number Re during the pre-quarantine period at ~3.7, which is rather high. Multiple superspreading events and the high Re can be due to some of the conditions specific to a hospital not specifically equipped for infection control, including dense contacts (in particular, spread by medical workers), absence of protective measures, and lack of awareness. In the second phase of the outbreak, we observe a significant decrease in Re down to ~1.4. This change can be explained by two factors. Firstly, it can be due to increased awareness and quarantine measures which were in effect after April 7. Secondly, it can be due to a large number of people already ill, preventing further infection; indeed, around 30% of people at the hospital had been infected by April 22. We cannot quantify the contribution of these factors to the slowing rate of infection spread with available data and methods.

The Vreden hospital outbreak has apparently contributed to COVID-19 spread in Saint Petersburg. By May 26, more than 14,000 cases had been confirmed in Saint Petersburg. In our dataset, the three predominantly-Vreden groups include five non-Vreden samples (out of the 84 non-Vreden Saint Petersburg samples; Fig. 2 and Supplementary Fig. 3), indicating transmission of SARS-CoV-2 from the hospital to the outside population. For two of these samples, 6840 and 6846, their relation to the Vreden hospital could be identified: these two cases were probably infected by family members of a Vreden hospital employee. As we didn’t specifically target Vreden-related samples collected outside of the Vreden hospital, the high proportion of Vreden-related samples (5/84) implies that the Vreden hospital outbreak has conceivably contributed to a substantial fraction of non-Vreden cases in Saint Petersburg.

## Methods

### Sample collection and sequencing

Nasopharyngeal and/or throat swabs were collected in virus transport media. Total RNA was extracted using RiboPrep DNA/RNA extraction kit (AmpliSens, Russia). Extracted RNA was immediately tested for SARS-CoV-2 using LightMix® SarbecoV E-gene plus EAV control (TIB Molbiol, Berlin, Germany) provided by the WHO Regional Office for Europe and based on *Charite* protocol ^65^. LightMix® SarbecoV E-gene plus EAV control was used with BioMaster qRT-PCR Kit (Biolabmix, Russia). Briefly, each 20 μL reaction mixture contained 10 μL of 2x buffer, 0,5 μL of LightMix SarbecoV E-gene reagent mix, 4.7 μL of nuclease-free water, 0.8 μL of enzyme, and 4 μL of extracted RNA as the template. RT-PCR was performed on a LightCycler 96 RT-PCR system (Roche). The thermal cycling conditions were 55 °C for 15 min, 95 °C for 5 min, followed by 45 cycles of 95 °C for 5 s, 60 °C for 15 s and 72 °C for 15 s.

Specimens with Ct values less than 30 were selected for whole-genome sequencing.

### Virus isolation

For some of the SARS-CoV-2 PCR-positive samples (see Supplementary Data 2), viruses were isolated in Vero cell culture (ATCC #CCL-81). Cells were propagated in MEM (Gibco) supplemented with GlutaMax (Gibco), Sodium Pyruvate (Gibco) and 10% FBS (Gibco #10500). 2 days before inoculation cells were seeded in 5.5 cm2 cell culture tubes (Nunc) at 1:4 ratio and 5% FBS. Samples were diluted 1:10 with serum free media containing antibiotic-antimycotic (Gibco) and inoculated to cells in a volume 0.5 ml/tube. After incubation for 2 h at 37C, inoculum was removed and 3 ml of serum free media with anti-anti was added to tubes. Viruses were harvested 4-6 days post inoculation (p.i.) when cytopathic effect (CPE) was near 80-100%, while first signs of CPE were typically observed 2-4 days p.i. For subsequent work, 0.15 ml of virus suspension was lysed in 0.5 ml RLT buffer (QIAGEN) and stored at −20C until RNA extraction.

### Whole-genome sequencing

RNA from primary clinical specimens and virus isolates was re-extracted using QIAamp Viral RNA Mini Kit or RNeasy Mini Kit (QIAGEN). Whole-genome amplification of SARS-CoV-2 virus genome was performed using ARTIC Network protocol ^66^ with modifications. ARTIC Network primer sets were modified by adding ONT universal tags: 5’-TTTCTGTTGGTGCTGATATTGC-3’ and 5’-ACTTGCCTGTCGCTCTATCTTC-3’ for forward and reverse primers, respectively (see Supplementary Data 2 for details). 1D Ligation sequencing kit (SQK-LSK109) with PCR barcoding expansion (EXP-PBC096) was utilized for sequencing library preparation. MinION (Oxford Nanopore) (flow cell R9.4.1) was used for whole-genome sequencing.

### Genome assembly and consensus correction

Fast5 files produced by minION were basecalled using guppy_basecaller v3.6.0 ^67^. Basecalled reads were processed by Porechop ^68^ in two steps. First, for each sequencing run, reads were demultiplexed with default settings, with built-in barcode and adapter sequences cleaved from read ends. Second, PCR primers were trimmed from demultiplexed reads with options --end_size 70 --no_split. Processed reads corresponding to one sample were combined.

For each sample, we then mapped reads onto the Wuhan-Hu-1 SARS-CoV-2 genome sequence (NCBI ID: MN908947.3) using minimap2 ^69^ with default settings and filtered out chimeric reads and reads that had secondary alignments. SAMtools-mpileup ^70^ was used to produce draft consensus sequences which were then corrected as follows. Mappings were converted into .tsv files using sam2tsv ^71^, and for each position in the genome, we computed the frequencies of all variants present. We further considered positions with coverage 15 or higher and alternative (compared to Wuhan-Hu-1) variant frequency 50% or higher. We corrected the draft consensus sequences based on the defined set of alternative variants. Each introduced correction was assessed by visually analyzing the corresponding region of mapped reads in IGV ^72^. Additionally, we manually assessed all alternative variants that had coverage 100 and below. We observed several spurious mutations that were not included in final consensus sequences, including the homoplasic mutation G11083T residing at the end of the poly-T tract in the genome that was observed in five of our samples.

### SARS-CoV-2 dataset preparation and filtering

All complete high coverage genomes of SARS-CoV-2 for all regions were downloaded from GISAID on May 26, 2020, for a total of 20469 global sequences and 78 Russian sequences (Supplementary Data 1). To this dataset we added the 136 sequences obtained in this study. Sequences shorter than 29,000 bp, sequences with more than 300 positions with missing data (Ns), sequences excluded by Nextstrain, and samples corresponding to resequencing of the same patients were removed. This led to exclusion of one Russian sample sequenced in this study (hCoV-19/Russia/Ulan-Ude-RII4560S/2020), as well as 834 non-Russian sequences (Supplementary Data 3).

The obtained sequences were aligned with MAFFT 7.453 ^73^ with the following parameters: ‘ --addfragments --keeplength’. As the reference sequence, we utilized Wuhan-Hu-1/2019 (NCBI ID: MN908947.3). To remove low-quality bases from the alignment, 100 nucleotides from the beginning and from the end were trimmed. The final alignment was used to construct the phylogenetic tree with IQ-Tree 1.6.12 ^74^ with GTR substitution model and ‘-fast’ option. We used TreeTime ^75^ to reconstruct the sequences of the internal tree nodes. Sequences separated from the tree root by more than ten nucleotide mutations were excluded as probable results of incorrect base calling; this included two sequences from Russia (Russia/SCPM-O-02/2020 and Russia/SCPM-O-05/2020) (Supplementary Data 3).

The final dataset contained 19834 virus SARS-CoV-2 sequences. The resulting tree is available as Supplementary Fig. 1.

### Phylogenetic analysis

We categorized each Russian sequence into one of the five phylogenetic categories based on its phylogenetic position, defined as follows. A Russian transmission lineage is a set of two or more sequences that form a Russian-only clade. A Russian singleton is a single Russian sequence that forms a clade of its own (i.e., possesses one or more private mutations) and is not a part of a Russian transmission lineage. A Russian stem cluster is a set of Russian sequences identical to each other and to some non-Russian sequences. A Russian stem-derived transmission lineage is a Russian transmission lineage whose immediate ancestor is a Russian stem cluster, and at least one of the sequences in this stem cluster was collected earlier than the earliest sequence in the lineage. A Russian stem-derived singleton is a Russian singleton whose immediate ancestor is a Russian stem cluster, and at least one of the sequences in this stem cluster was collected earlier than the singleton. These categories are schematically represented on Supplementary Figure 2.

As sampling dates, we used the collection dates reported in GISAID. For some samples, either the day or both the day and the month of collection were missing. In such cases, the date was set to the latest date possible, e.g. “2020-03” to “2020-03-31” and “2020” to “2020-12-31”. The date for the lineage introduction was estimated as the earliest collection date among all samples in the particular lineage.

In phylogeographic analysis, we assumed that the possible source(-s) of introduction for Russian transmission lineages and Russian singletons were the sampling country(-ies) of the non-Russian sequences ancestral to the considered lineage; for Russian stem-derived transmission lineages and Russian stem-derived singletons, these were the non-Russian sequences identical to the ancestral stem cluster. For stem clusters, the possible source of introduction were the countries of origin for the sequences identical to those in the stem cluster. As a possible source of introduction, we only considered those countries with the earliest collection date earlier than the earliest collection date among all samples in the lineage (or than the collection date of the singleton). For patients with known travel history, we considered the country (continent) of origin as uniquely identified by phylogeography if it was either the only country (continent) on the ancestral stem, or the one with the earliest collection date. For samples with no travel data, the same logic was applied, except we only infer the country or continent of origin if it was the only one on the stem. If there were more than eight countries in the list, countries were merged into regions: Africa, Asia, Europe, North America and South America. To study the possibility of introduction from China, we performed the same analysis, but considered China, Hong Kong and Taiwan separately from the rest of Asia.

To estimate the number of possible exports out of Russia, we used maximum parsimony, asking whether any non-Russian sequences were nested within Russian clades. In this analysis, we conservatively assumed that the phylogenetic nodes carrying both Russian and non-Russian sequences were positioned outside Russia. No evidence for export was observed.

To understand whether introductions to Russia occurred through major transportation hubs (Moscow and Saint Petersburg), we considered all Russian samples not included in Russian transmission lineages. For these samples, we calculated the branch lengths from each sample to its immediate ancestor, and labeled all samples by two categories: major hubs (Moscow, Moscow region, Saint Petersburg and Leningrad region) and other locations (samples from all other locations in Russia). On this data, we performed a permutation test, shuffling labels across the dataset 1000 times. The two-sided p-value was calculated.

### Phylodynamics of SARS-CoV-2 in Vreden hospital

As discussed in the Results, the Vreden samples belong to three distinct phylogenetic groups. To account for this, we constrained the phylogeny as follows: (((group1), group 2),(group 3)). We independently ran BEAST2 on three datasets: (i) the whole Vreden dataset comprising groups 1, 2 and 3; (ii) groups 1 and 2; and (iii) group 1 only. The model details were as follows. The effective reproductive number was allowed to change on March 27 (which delimits the suspected out-of-hospital period) and again on April 8 (which corresponds to the introduction of quarantine). The sampling proportion was set to zero prior to March 27, and was allowed to change on April 8 and April 15. The prior on the clock rate was set to be a normal distribution with mean 9.41*10^−4^ and standard deviation 4.99*10^−5^, based on the estimates from the UK study ^47^. Dates were fixed for the 11 sequences (all from group 1) for which the dates of symptom onset were available; for the remaining 41 sequences, we applied tip date sampling with a uniform prior as detailed below. Other priors are provided in Supplementary Table 7. Supplementary Tables 4, 5 and 6 contain the Bayesian estimates of the model parameters for three datasets comprising groups 1, 2 and 3, groups 1 and 2 and group 1, respectively.

The fifty two Vreden samples were collected on 5 distinct dates, with a substantial lag between subsequent collection dates (see Supplementary Table 3). For some of the samples, sample collection date could differ substantially from the symptoms onset date. To address the bias in collection dates, we used the symptoms onset date instead of the collection date in BEAST2 analysis, estimating it as follows. For 11 samples, the symptoms onset dates were known (Supplementary Table 2). For each of the remaining 41 samples, we produced a posterior estimate of its symptoms onset date by using a uniform prior between March 31st and the collection date.

### Public information and data visualization

The initial Russian map was downloaded from GADM (sf, level 1) ^76^. Numbers of confirmed cases in Russia by region were downloaded on May, 26, 2020 from ref. ^77^. Patients age data for Russian samples were extracted from GISAID metadata; for 12 samples, age data were missing. The Spearman correlation between age and collection date was calculated in R version 3.6.3 with cor.test() function. Maps were visualized with the ggplot2 package in R. Phylogenetic trees were visualized with the ETE3 toolkit ^78^ in Python 3 and iTOL v4 ^79^. The maximum clade credibility tree was visualized with FigTree ^80^.

## Data availability

GISAID accession IDs of SARS-CoV-2 consensus sequences produced in this study are provided in Supplementary Data 4. The results of minION Nanopore whole-genome sequencing were deposited in SRA (accession numbers pending; will be available by BioProject ID PRJNA645970). Description for all supplementary files is summarized in Supplementary Information.

## Code availability

Python/R scripts used for data visualization are available upon request.

## Data Availability

GISAID accession IDs of SARS-CoV-2 consensus sequences produced in this study are provided in Supplementary Data 4. The results of minION Nanopore whole-genome sequencing were deposited in SRA (accession numbers pending; will be available by BioProject ID PRJNA645970).

## Acknowledgements

We thank Dr. Josh Quick and Dr. Nick Loman (University of Birmingham) for providing the ARTIC Network primer set version 2 for whole-genome amplification of SARS-CoV-2 viruses; Prof. Rasmus Nielsen for his insights in setting up the BEAST2 analysis; Dr. Carlo Pacioni for his help with setting tip dates sampling for BEAST2 analysis. We further thank colleagues from the Chumakov Federal Scientific Center for Research and Development of Immune- and Biological Products of Russian Academy of Sciences (Moscow, Russia) for their valuable help with BSL-3 sample processing. No compensation was received for their roles in the study. This research was supported in part through computational resources of HPC facilities at NRU HSE. N.S., V.Sp., D.G. and V.S. performed the research within the framework of the HSE University Basic Research Program. We thank all of the authors who have contributed genome data on GISAID (see Supplementary Data 1 for the list).

## Author contributions

A.B.K., A.V.F., M.V.S., A.A.I., D.M.D. and D.L. performed RT-PCR testing, isolated the virus, collected, sequenced, assembled and curated the viral genomes. K.R.S. and A.V.F. performed base calling. K.R.S. and S.K.G. analyzed genomic data and performed phylogenetic analyses. G.A.B. and S.K.G. analyzed geographic data, travel data and GISAID metadata. O.V.S. collected patient samples at the Vreden hospital and contributed to detailed epidemiological analysis. K.R.S, N.S., V.Sp., D.G. and V.S prepared a phylodynamics pipeline, explored probabilistic models and their limitations for the study. N.S. and V.S. performed birth-deaths skyline analysis of the Vreden hospital outbreak. G.A.B. conceived, designed and supervised the project and drafted the manuscript. K.R.S., S.K.G., V.S. and G.A.B. wrote the manuscript with contributions from all authors. All authors read the manuscript and agree to its contents.

## References

1. Coronavirus Update (Live): 11,965,661 Cases and 546,988 Deaths from COVID-19 Virus Pandemic - Worldometer. https://www.worldometers.info/coronavirus/#countries.

2. Coronavirus disease 2019 (COVID-19): Situation report, 42. (2020).

3. Coronavirus disease 2019 (COVID-19): Situation report, 47. (2020).

4. Coronavirus disease 2019 (COVID-19) : Situation report, 53. (2020).

5. The Moscow Times. Russia Closes Far East Border Over Coronavirus. https://www.themoscowtimes.com/2020/01/30/russia-closes-far-east-border-over-coronavirus-a69100 (2020).

6. Russia restricts air travel with China from February 1 due to coronavirus. TASS https://tass.com/economy/1115335.

7. In Russian. Прнорешиео временнӣ ӣграничении движения через пункты пропуска на отдельных участках государственной границы Российской Федерации с Китайской Народной Республикой. http://government.ru/docs/38879.

8. In Russian. Принят ряд решений в целях предупреждения проникновения на территорию России коронавирусной инфекции с территории Китайской Народной Республики. http://government.ru/docs/38900/.

9. In Russian. Принято решение о временном ограничении въезда граждан иностранных государств с территории Китайской Народной Республики в воздушных пунктах пропуска через государственную границу Российской Федерации. http://government.ru/docs/38912/.

10. In Russian. Принято решение о временной приостановке пропуска через государственную границу Российской Федерации граждан Китайской Народной Республики, въезжающих для осуществления трудовой деятельности, в частных, учебных и туристических целях. http://government.ru/docs/38996/.

11. In Russian. Принято решение о временном ограничении въезда иностранных граждан с территории Республики Корея в воздушных пунктах пропуска через государственную границу Российской Федерации. http://government.ru/docs/39041/.

12. In Russian. Временно ораничен въезд иностранных граждан с территории Исламской Республики Иран в воздушных пунктах пропуска через государственную границу Российской Федерации. http://government.ru/docs/39043.

13. Why are there so few reported COVID-19 cases in Russia? — Meduza. Meduza https://meduza.io/en/feature/2020/03/06/why-are-there-so-few-reported-covid-19-cases-in-russia.

14. First two persons infected with coronavirus identified in Russia. TASS https://tass.com/society/1115101.

15. One imported coronavirus case confirmed in Russia. TASS https://tass.com/society/1125627.

16. Covid-19: Global summary. Covid-19 https://epiforecasts.io/covid/posts/global/ (2020).

17. Coronavirus disease 2019 (COVID-19) : Situation report, 61. (2020).

18. In Russian. Объемы перевозок через аэропорты России. https://favt.ru/dejatelnost-ajeroporty-i-ajerodromy-osnovnie-proizvodstvennie-pokazateli-aeroportov-obyom-perevoz/.

19. In Russian. Статистика аэропорта Домодедово. https://business.dme.ru/company/finance/operacionnye-rezul_taty-/.

20. In Russian. Новости аэропорта Внуково. http://corp.vnukovo.ru/press/news/.

21. In Russian. Показатели аэропорта Пулково. https://pulkovoairport.ru/about/performance/.

22. In Russian. № 12-УМ от 05.03.2020 «О введении режима повышенной готовности». https://www.mos.ru/authority/documents/doc/43503220/.

23. In Russian. Коронавирус. Дополнительные меры 14.03.2020. https://www.sobyanin.ru/koronavirus-dopolnitelnye-mery-14-03-2020.

24. In Russian. Коронавирус. Запрет проведения массовых мероприятий и другие ограничительные меры 16.03.2020. https://www.sobyanin.ru/koronavirus-ogranichitelnye-mery-16-03-2020.

25. In Russian. Постановление правительства Санкт-Петербурга от 13 марта 2020 года № 121 ‘О мерах по противодействию распространению в Санкт-Петербурге новой коронавирусной инфекции (COVID-19)’. Российская газета.

26. In Russian. Принято решение о временной приостановке пропуска через государственную границу Российской Федерации иностранных граждан и лиц без гражданства, прибывающих с территории Итальянской Республики для обучения и трудовой деятельности, а также в частных, туристических и транзитных целях. http://government.ru/docs/39140/.

27. In Russian. Принято решение о временном ограничении въезда в Российскую Федерацию иностранных граждан и лиц без гражданства, в том числе прибывающих с территории Республики Беларусь, а также граждан Республики Беларусь. http://government.ru/docs/39179/.

28. In Russian. Объемы перевозок через аэропорты МАУ за январь-апрель 2020 года. http://www.aex.ru/docs/2/2020/5/29/3074/.

29. In Russian. Пассажиропоток аэропорта ‘Шереметьево’ в мае возрос на 39%. http://www.aex.ru/news/2020/6/23/213922/.

30. Coronavirus disease 2019 (COVID-19) : Situation report, 51.

31. Coronavirus disease 2019 (COVID-19) : Situation report, 94.

32. GISAID - Initiative. https://www.gisaid.org/.

33. Rambaut, A. et al. A dynamic nomenclature proposal for SARS-CoV-2 to assist genomic epidemiology. bioRxiv 2020.04.17.046086 (2020) doi:10.1101/2020.04.17.046086.

34. GISAID - Next hCoV-19 App. https://www.gisaid.org/epiflu-applications/next-hcov-19-app/.

35. Initial tests showed coronavirus in five more people in Yakutia. https://webcache.googleusercontent.com/search?q=cache:yVHzr1z2I6kJ: https://www.corona24.news/c/2020/03/19/initial-tests-showed-coronavirus-in-five-more-people-in-yakutia-society-rbc.html+&cd=3&hl=en&ct=clnk&gl=ca.

36. In Russian. Главу газовой компании Якутии госпитализировали из-за подозрения на вирус. РБК https://www.rbc.ru/society/18/03/2020/5e71fc479a7947187644b347.

37. Two residents of Chechnya infected with coronavirus during Hajj. Caucasian Knot https://www.eng.kavkaz-uzel.eu/articles/50388/.

38. Most of Moscow’s New Coronavirus Patients Younger Than 40. https://www.themoscowtimes.com/2020/03/30/most-of-moscows-new-coronavirus-patients-younger-than-40-a69797 (2020).

39. nextstrain.org/ncov. https://nextstrain.org/ncov/.

40. In Russian. Война с коронавирусом в замкнутом пространстве. История закрытой на карантин клиники в Петербурге. BBC News Русская служба https://www.bbc.com/russian/features-52813191 (2020).

41. Russian doctors, nurses face more risks as virus cases grow. AP NEWS https://apnews.com/b4950726aea5b4ec33a0f4d8fa76cb40 (2020).

42. Stadler, T., Kühnert, D., Bonhoeffer, S. & Drummond, A. J. Birth-death skyline plot reveals temporal changes of epidemic spread in HIV and hepatitis C virus (HCV). Proc. Natl. Acad. Sci. U. S. A. 110, 228–233 (2013).

43. Bouckaert, R. et al. BEAST 2.5: An advanced software platform for Bayesian evolutionary analysis. PLoS Comput. Biol. 15, e1006650 (2019).

44. arambaut et al. Phylodynamic Analysis | 176 genomes | 6 Mar 2020. Virological https://virological.org/t/phylodynamic-analysis-176-genomes-6-mar-2020/356 (x2020).

45. da Silva Candido, D. et al. Evolution and epidemic spread of SARS-CoV-2 in Brazil. medRxiv 2020.06.11.20128249 (2020).

46. lemieux. Introduction and spread of SARS-CoV-2 in the greater Boston area. Virological https://virological.org/t/introduction-and-spread-of-sars-cov-2-in-the-greater-boston-area/503 (2020).

47. OliverPybus, Kristian_Andersen ljones & gbellobr. Preliminary analysis of SARS-CoV-2 importation & establishment of UK transmission lineages. Virological https://virological.org/t/preliminary-analysis-of-sars-cov-2-importation-establishment-of-uk-transmission-lineages/507/2 (2020).

48. ggithinji. Introduction and local transmission of SARS-CoV-2 cases in Kenya. Virological https://virological.org/t/introduction-and-local-transmission-of-sars-cov-2-cases-in-kenya/497 (2020).

49. Miller, D. et al. Full genome viral sequences inform patterns of SARS-CoV-2 spread into and within Israel. medRxiv 2020.05.21.20104521 (2020).

50. Genomic Epidemiology of SARS-CoV-2 in Guangdong Province, China. Cell 181, 997–1003.e9 (2020).

51. Grubaugh, N. D. et al. Genomic epidemiology reveals multiple introductions of Zika virus into the United States. Nature 546, 401–405 (2017).

52. Kraemer, M. U. G. et al. Reconstruction and prediction of viral disease epidemics. Epidemiol. Infect. 147, (2019).

53. Gonzalez-Reiche, A. S. et al. Introductions and early spread of SARS-CoV-2 in the New York City area. Science (2020) doi:10.1126/science.abc1917.

54. Villabona-Arenas, C. J., Hanage, W. P. & Tully, D. C. Phylogenetic interpretation during outbreaks requires caution. Nature Microbiology 5, 876–877 (2020).

55. Olsen, S. J. et al. Early Introduction of Severe Acute Respiratory Syndrome Coronavirus 2 into Europe. Emerg. Infect. Dis. 26, 1567–1570 (2020).

56. Böhmer, M. M. et al. Investigation of a COVID-19 outbreak in Germany resulting from a single travel-associated primary case: a case series. The Lancet Infectious Diseases (2020) doi:10.1016/s1473-3099(20)30314-5.

57. Evidence for Limited Early Spread of COVID-19 Within the United States, January–February 2020. MMWR Morb. Mortal. Wkly. Rep. 69, (2020).

58. Nadeau, S. A., Vaughan, T. G., Sciré, J., Huisman, J. S. & Stadler, T. The origin and early spread of SARS-CoV-2 in Europe. medRxiv 2020.06.10.20127738 (2020).

59. Stadler, T. Phylodynamic Analyses of outbreaks in China, Italy, Washington State (USA), and the Diamond Princess. Virological https://virological.org/t/phylodynamic-analyses-of-outbreaks-in-china-italy-washington-state-usa-and-the-diamond-princess/439 (2020).

60. Mizumoto, K., Kagaya, K., Zarebski, A. & Chowell, G. Estimating the asymptomatic proportion of coronavirus disease 2019 (COVID-19) cases on board the Diamond Princess cruise ship, Yokohama, Japan, 2020. Eurosurveillance 25, 2000180 (2020).

61. Estimation of the reproductive number of novel coronavirus (COVID-19) and the probable outbreak size on the Diamond Princess cruise ship: A data-driven analysis. Int. J. Infect. Dis. 93, 201–204 (2020).

62. Sekizuka, T. et al. Haplotype networks of SARS-CoV-2 infections in the Diamond Princess cruise ship outbreak. medRxiv 2020.03.23.20041970 (2020).

63. Deng, X. et al. Genomic surveillance reveals multiple introductions of SARS-CoV-2 into Northern California. Science (2020) doi:10.1126/science.abb9263.

64. Report into a nosocomial outbreak of coronavirus disease 2019 (COVID-19) at Netcare St. Augustine’s Hospital. https://www.krisp.org.za/manuscripts/StAugustinesHospitalOutbreakInvestigation_Final Report_15may2020_comp.pdf.

65. Corman, V. M. et al. Detection of 2019 novel coronavirus (2019-nCoV) by real-time RT-PCR. Eurosurveillance 25, 2000045 (2020).

66. joshquick.joshquick/artic-ncov2019. https://github.com/joshquick/artic-ncov2019.

67. Oxford Nanopore Technologies. https://github.com/nanoporetech.

68. Porechop. https://github.com/rrwick/Porechop.

69. Li, H. Minimap2: pairwise alignment for nucleotide sequences. Bioinformatics 34, 3094–3100 (2018).

70. Li, H. A statistical framework for SNP calling, mutation discovery, association mapping and population genetical parameter estimation from sequencing data. Bioinformatics 27, 2987–2993 (2011).

71. Jvarkit : Java utilities for Bioinformatics. http://lindenb.github.io/jvarkit/.

72. Robinson, P. & Jtel, T. Z. Integrative genomics viewer (IGV): Visualizing alignments and variants. Computational Exome and Genome Analysis 233–245 (2017) doi:10.1201/9781315154770-17.

73. Katoh, K. & Standley, D. M. MAFFT multiple sequence alignment software version 7: improvements in performance and usability. Mol. Biol. Evol. 30, 772–780 (2013).

74. Nguyen, L.-T., Schmidt, H. A., von Haeseler, A. & Minh, B. Q. IQ-TREE: a fast and effective stochastic algorithm for estimating maximum-likelihood phylogenies. Mol. Biol. Evol. 32, 268–274 (2015).

75. Sagulenko, P., Puller, V. & Neher, R. A. TreeTime: Maximum-likelihood phylodynamic analysis. Virus Evol 4, vex042 (2018).

76. GADM. https://gadm.org/download_country_v3.html.

77. In Russian. Оперативные данные. https://xn--80aesfpebagmfblc0a.xn--p1ai/information/.

78. Huerta-Cepas, J., Serra, F. & Bork, P. ETE 3: Reconstruction, Analysis, and Visualization of Phylogenomic Data. Mol. Biol. Evol. 33, 1635–1638 (2016).

79. Letunic, I. & Bork, P. Interactive Tree Of Life (iTOL) v4: recent updates and new developments. Nucleic Acids Res. 47, W256–W259 (2019).

80. FigTree. http://tree.bio.ed.ac.uk/software/figtree/.

